# Effects of 4-Month Methylphenidate Treatment on Functional Connectivity in Male Individuals with Attention-Deficit Hyperactivity Disorder

**DOI:** 10.1101/2025.08.27.25334547

**Authors:** Eline Vansina, Linda Douw, Antonia Kaiser, Zarah van der Pal, Daphne E. Boucherie, Taco J. De Vries, Tommy Pattij, Jessica R. Cohen, Pieter J. Hoekstra, Liesbeth Reneman, Anouk Schrantee

## Abstract

**Objective:** Methylphenidate is effective in reducing ADHD symptoms in the short term, but long-term benefits are inconsistent, possibly due to the development of tolerance. Moreover, little is known about its sustained effects on brain functional connectivity. We examined whether a 4-month methylphenidate treatment leads to sustained alterations in resting-state functional connectivity, and whether acute brain responses to methylphenidate decrease after treatment, as a potential marker of neurobiological tolerance.

**Method:** This is a secondary analysis of the ePOD-MPH RCT in which 50 boys and 49 men with ADHD were randomized to methylphenidate or placebo. Resting-state fMRI data were collected before, and one week after, a 4-month treatment period. At both visits, participants were scanned before and after an acute oral methylphenidate challenge. We computed whole-brain and default mode network (DMN) global efficiency, and DMN–whole-brain connectivity strength.

**Results:** In adults, methylphenidate (but not placebo) led to sustained increases in whole-brain efficiency (p<0.01) and DMN–whole-brain connectivity strength (p=0.03). No significant effects were observed in children (all p’s>0.17). Exploratory analyses indicated that whole-brain efficiency increases related to decreasing cognitive performance in methylphenidate-treated children, but improving performance in placebo-treated children, suggesting treatment-dependent moderation (p<0.01). Acute connectivity responses remained stable in adults across visits (all p’s>0.15), but increased in children, regardless of treatment (all p’s<0.04).

**Conclusion:** Four months of methylphenidate treatment led to sustained functional connectivity changes in adults with ADHD, without evidence of neurobiological tolerance. These findings emphasize the importance of studying longer treatment durations, considering that methylphenidate treatments typically span multiple years.

## Introduction

Attention-deficit hyperactivity disorder (ADHD) is a neurodevelopmental disorder characterized by age-inappropriate and persistent levels of inattention, hyperactivity and impulsivity, negatively impacting quality of life. Methylphenidate, also known under the brand name Ritalin, is commonly used as the first-line pharmacological treatment for ADHD. In the short term, methylphenidate treatment has been shown to have positive effects on ADHD symptoms, reducing hyperactivity, impulsivity, and inattention[1].

This is in contrast with the lack of evidence for the long-term efficacy of the drug. The Multimodal Treatment study of ADHD found that none of the initial treatment benefits of methylphenidate were maintained in the long term, despite medication doses substantially increasing over time[2]. Moreover, a placebo-controlled discontinuation study found that 60% of participants did not experience worsening of symptoms following discontinuation of treatment[3]. Although the long-term efficacy of methylphenidate remains insufficiently studied, evidence for sustained treatment benefits beyond two years is limited[4-6]. Nonetheless, some studies indicate that continued treatment may be associated with reduced criminal behavior and improved educational outcomes[7,8].

Despite this, the neurobiological effects of prolonged methylphenidate use remain poorly understood, including the timing at which such changes may emerge. Existing evidence suggests that use of methylphenidate in childhood and adolescence may impact the development of brain structure and function. For example, animal studies have shown age-dependent structural and functional changes in frontostriatal networks[9,10] and striatal dopamine transporter (DAT) upregulation following pre-pubescent exposure to methylphenidate[11]. Similarly, DAT upregulation has been observed in adults with ADHD following twelve months of methylphenidate treatment[12]. Furthermore, blood flow responses to a methylphenidate challenge were altered in the striatum and thalamus of children, but not adults, up to one week after a 4-month treatment period[13].

Some of these neurobiological changes may contribute to the phenomenon of *neurobiological tolerance;* the development of adaptive changes in the brain that reduce responsiveness to acute drug administration after prolonged use[14]. The development of neurobiological tolerance to methylphenidate may explain the reduced long-term efficacy of methylphenidate. Supporting this, human and preclinical studies indicate the development of both acute[15] and long-term[16-18] behavioral tolerance, although the underlying neurobiological mechanisms and the interaction with brain development remain unclear.

Given that ADHD is increasingly conceptualized as a disorder of large-scale brain dysconnectivity, it is highly relevant to investigate how methylphenidate affects functional connectivity. Evidence from meta-analyses has consistently shown atypical connectivity within and between the default mode network (DMN), frontoparietal network, and attention networks in individuals with ADHD[19,20]. Evidence for the role of altered connectivity of the DMN within the connectome is particularly prominent[20].The DMN supports internally directed processes such as mind-wandering and self-reflection, and its dysregulation has been linked to cognitive and behavioral difficulties in ADHD[20]. Acute administration of methylphenidate has been found to normalize functional connectivity of the DMN, insula, and striatum in people with ADHD, i.e., more in line with typically developing controls[21]. Moreover, we previously found, in the same cohort as studied here, that an acute methylphenidate challenge induced distinct functional connectivity changes in children and adults, suggesting age-dependent acute effects[22]. Task-fMRI studies have also demonstrated acute methylphenidate-induced functional connectivity increases, which were associated with improvements in reaction time[23], working memory[24], and inhibitory control[25].

As of yet, research on the long-term effects of methylphenidate on functional connectivity is scarce. In children, increases in functional connectivity have been reported between the DMN and the putamen after six months of treatment with methylphenidate, which correlated with the reduction of ADHD-related symptom severity[26]. In line with these findings, one study observed that normalization of functional connectivity of the right inferior gyrus and the bilateral cerebellum correlated with improvements in clinical symptoms following three months of methylphenidate treatment in children[27]. In another study, poor treatment response was associated with functional connectivity increases within the cingulo-opercular network, whereas treatment responders showed stable functional connectivity patterns over time[28]. These findings suggest that long-term methylphenidate treatment is associated with changes in functional connectivity, but whether these changes sustain beyond treatment-end remains unclear.

Therefore, this study investigated whether a 4-month methylphenidate treatment led to sustained alterations in functional connectivity after a 1-week washout, and whether these alterations were linked to changes in the cognitive domains of response inhibition and psychomotor speed. We hypothesized to find sustained alterations in functional connectivity, measured in terms of network efficiency and connectivity strength, across the whole brain and the DMN. Moreover, to investigate whether neurobiological tolerance develops after a 4-month treatment, we assessed alterations in functional connectivity (i.e. *connectivity response*) to an acute administration (*challenge*) of methylphenidate. We expected reduced connectivity responses to an acute methylphenidate challenge after a 4-month treatment. Lastly, we investigated to what extent these changes are age-dependent by qualitatively comparing results in male children and adults. We expected these effects to be more pronounced in children than in adults, due to the ongoing development of the dopaminergic system in children.

## Methods

### Participants

This is a secondary analysis of the double-blind, placebo controlled “effects of Psychotropic drugs On the Developing brain - methylphenidate” (ePOD-MPH) trial[29]. Fifty treatment-naive boys (aged 10–12 years) and 49 treatment-naive men (aged 23– 40 years), diagnosed with ADHD and in need of pharmacotherapy, were recruited for the trial. The primary objective of the ePOD-MPH trial was to investigate the effects of methylphenidate on the dopaminergic system, and the modulation by age, assessed using pharmacological MRI[13]. Here we report on a secondary analysis focusing on rs-fMRI. Participants were recruited through clinical programs at the Child and Adolescent Psychiatry Center Triversum (Alkmaar, The Netherlands), the Department of Child and Adolescent Psychiatry at the Bascule/AMC (Amsterdam, The Netherlands), and the PsyQ Mental Health Facility (The Hague, The Netherlands). For more information on the participant inclusion and exclusion criteria, see Supplementary Materials.

### Study design

Participants received four months of treatment with either methylphenidate or placebo, followed by a 1-week washout period, during which participants were instructed not to take any methylphenidate (Figure 1A). This wash-out period duration was chosen to fully eliminate the acute pharmacological effects of methylphenidate during the follow-up scan. Before the start of the treatment (i.e., baseline) and following washout (i.e., follow-up), participants underwent two resting-state functional MRI (rs-fMRI) scans: before and 90 minutes after an acute methylphenidate challenge (oral dose of short-acting methylphenidate; 0.5mg/kg with a maximum of 20mg in children and 40mg in adults; Sandoz B.V., Weesp, the Netherlands). In addition, behavioral and cognitive data were collected (Supplementary Materials).

**Fig 1.**
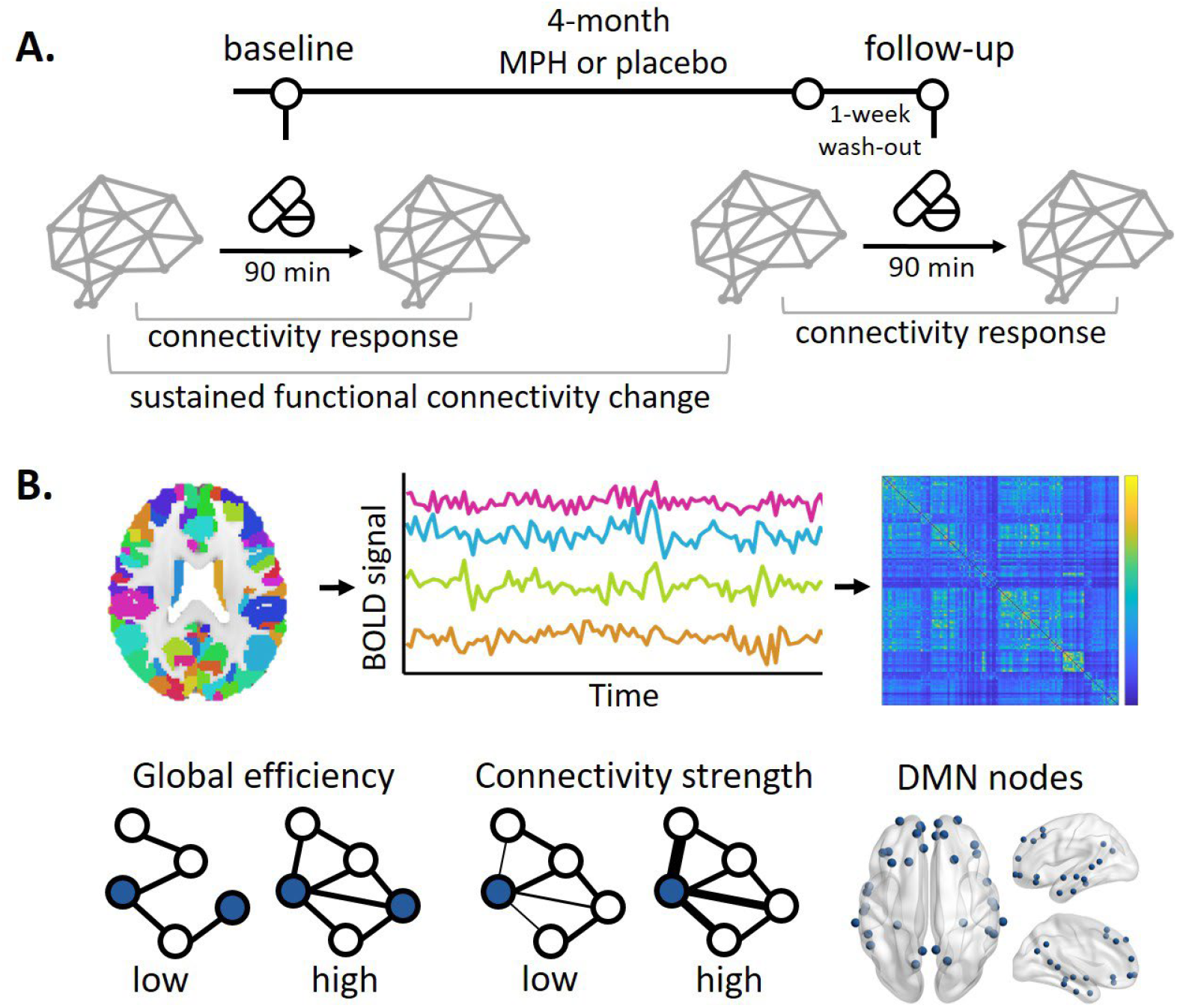
Study design and analysis overview. **a)** Study design. **b)** Analysis overview. DMN = Default Mode Network; BOLD = Blood-oxygen-level-dependent; MPH = methylphenidate

### MRI acquisition and processing

MR scans were made on 3T Philips scanners (Philips Healthcare, Best, The Netherlands) using an 8-channel receive-only head coil. A 3D T1-weighted anatomical scan (TR/TE = 9.8/4.6 ms, FOV = 256 × 256 × 120, voxel size = 0.875×0.875×1.2 mm) was acquired for registration purposes, and rs-fMRI scans were acquired using a single-shot echo-planar imaging sequence (TR/TE = 2300/30 ms, resolution = 2.3 × 2.3 × 3 mm, 39 sequential slices, FOV = 220 x 220 × 117 mm, GE-EPI read-out, no gap, FA = 80°, volumes = 130).

The rs-fMRI data were preprocessed using fMRIPrep v1.2.3[30], and processing included ICA-AROMA denoising (Supplementary Materials). Subsequently, white matter and cerebrospinal fluid signals, obtained before ICA-AROMA, were regressed from the signal, and a low-pass filter (100s) was applied using FSL. Framewise displacement (FD) values were calculated from the low-pass filtered time series[31]. The brain was parcellated into 246 nodes using the Brainnetome atlas[32], and time series per region were extracted per scan. High-motion volumes (FD>0.3 mm), including the volume preceding and following the high-motion volume, were scrubbed from the time series. Scans were excluded from analysis if fewer than 104 volumes remained after scrubbing. Connectivity matrices were then constructed by calculating the absolutized Pearson correlation coefficient between each node’s timeseries, z-scoring the matrix, and rescaling the z-scores to between 0 and 1. Thirty-four low temporal signal-to-noise (tSNR) nodes were excluded from analysis, including nodes in occipital and temporal regions (Supplementary Materials).

Graph theory measures were calculated using the Brain Connectivity Toolbox[33] (v.2019-03-03) in Matlab (R2022b, Mathworks). For whole-brain metrics, all Brainnetome atlas nodes were included, except for low-tSNR nodes. The DMN was defined based on the Yeo 7-network parcellation[34] (Supplementary Materials). Global efficiency of the whole-brain (whole-brain efficiency) and within the DMN (DMN efficiency) were calculated as network measures of integration. Global efficiency is the inverse of the average shortest path lengths in a network and serves as a measure of information transmission efficiency. To assess connectivity between the DMN and the rest of the brain, connectivity strength was calculated. This reflects the importance of nodes in a network (wiring cost) and is calculated as the average sum of weights of the edges connected to a node. Higher weights of an edge indicate that two nodes in a network are strongly correlated. We averaged connectivity strength across all DMN nodes to result in a single estimate of connectivity strength between the DMN and the rest of the brain (DMN-whole-brain connectivity strength). To assess changes in connectivity responses to the acute methylphenidate challenge from baseline to follow-up, post-challenge network measures were subtracted from pre-challenge measures.

### Cognitive assessments

We assessed response inhibition and psychomotor speed, given their relevance to cognitive performance in ADHD. Response inhibition was measured with a go/no-go task (d-prime: hit rate minus false alarm rate), and psychomotor speed with a simple reaction time task (mean-RT and RT variability (SD-RT)). RT measures were inverted so that higher values reflect better performance. Task details are provided in the Supplementary Materials.

### Statistical analyses

*Main analysis* - Statistical analyses were performed using RStudio R4.4.1. Linear mixed models (LMM) were constructed with the network measures as the dependent variables (*lmerTest* package). Individual models were constructed for each age group (children, adults). The independent variables included in the model were visit (baseline, follow-up) and treatment group (methylphenidate, placebo). Statistical significance was evaluated using the Wald chi-square test (*car* package). Post-hoc FDR-corrected pairwise contrast analyses (*emmeans* package) were used to investigate the direction of the significant effects. To investigate the sustained effects of four months of methylphenidate treatment on functional connectivity, pre-challenge network measures were compared between baseline and follow-up. To investigate the connectivity response (i.e., development of tolerance) following a 4-month methylphenidate treatment, changes in network measures to the challenge were compared between baseline and follow-up. In addition, for each age group and visit, we tested whether the connectivity response differed from zero using paired t-tests.

*Exploratory analysis* - We conducted a multivariate analysis of variance (MANOVA, Pillai’s trace) to assess whether change in cognitive measures (i.e. across response inhibition and psychomotor speed) from baseline to follow-up differed as a function of changes in (pre-challenge) whole-brain efficiency and treatment group. For adults, we included d-prime, inverse mean-RT and inverse SD-RT as the outcome variables, whereas for children, d-prime was not included in the model due to significant data loss caused by a recording error. Additionally, we ran a follow-up analysis in adults excluding d-prime to allow better comparability with the model in children. Sensitivity analyses regarding the effect of motion are included in the Supplementary materials.

## Results

### Participant characteristics

One adult with ADHD was excluded from analysis due to undisclosed treatment with methylphenidate prior to the RCT. After excluding scans with excessive motion (defined as a mean FD>0.3), data from 42 children and 48 adults with ADHD were retained for analysis. Participant characteristics are summarized in Table 1 and the Supplementary Materials.

**Table 1.** Baseline characteristics of included participants. All values are presented as mean (SD) unless otherwise indicated. ^a^ For children: Wechsler Intelligence Scale for Children (WISC), for adults: National Adult Reading Test (NART)[35]. ^b^Disruptive Behavior Disorder Rating Scale. ^c^ ADHD - Self Report. IQ = intelligence quotient

|  | Children |  |  | Adults |  |  |
| --- | --- | --- | --- | --- | --- | --- |
|  | *Methylphenidate* | *Placebo* | *Statistic* | *Methylphenidate* | *Placebo* | *Statistic* |
|  | n=22 | n=20 |  | n=24 | n=24 |  |
| **Age (yrs)** | 11.39 (0.85) | 11.51 (0.10) | t(33.56)=-0.38, p=0.71 | 28.20 (4.38) | 29.00 (4.93) | t(45.38)=-0.60, p=0.55 |
| **IQ<sup>a</sup>** | 107.23 (20.83) | 102.28 (16.48) | t(37.97)=0.84, p=0.41 | 107.86 (8.75) | 107.86 (6.40) | t(38.47)=0.00, p=1.00 |
| **ADHD presentation (n)** |  |  |  |  |  |  |
| Inattentive | 12 | 10 |  | 11 | 5 |  |
| Hyperactive | 0 | 1 |  | 0 | 0 |  |
| Combined | 10 | 9 |  | 13 | 19 |  |
| **ADHD symptoms<sup>b,c</sup>** |  |  |  |  |  |  |
| DBD-RS Attention | 21.18 (3.07) | 22.80 (3.38) | t(38.53)=-1.62, p=0.11 | - | - |  |
| DBD-RS Hyperactivity | 14.73 (5.22) | 15.75 (6.92) | t(35.22)=-0.54, p=0.59 | - | - |  |
| ADHD-SR | - | - |  | 33.48 (9.78) | 32.13 (9.92) | t(41.74)=0.45, p=0.65 |
| **Compliance (%)** | 82.74 (15.53) | 77.45 (18.28) | t(33.01)=0.68, p=0.50 | 86.96 (7.97) | 85.71 (8.36) | t(47.77)=1.97, p=0.05 |
| **Methylphenidate challenge (mg)** | 17.78 (3.08) | 19.69 (1.25) | t(23.0)=-2.42, p=0.02 | 38.41 (2.38) | 38.04 (2.92) | t(42.0)=0.46, p=0.65 |

**Table 3.**
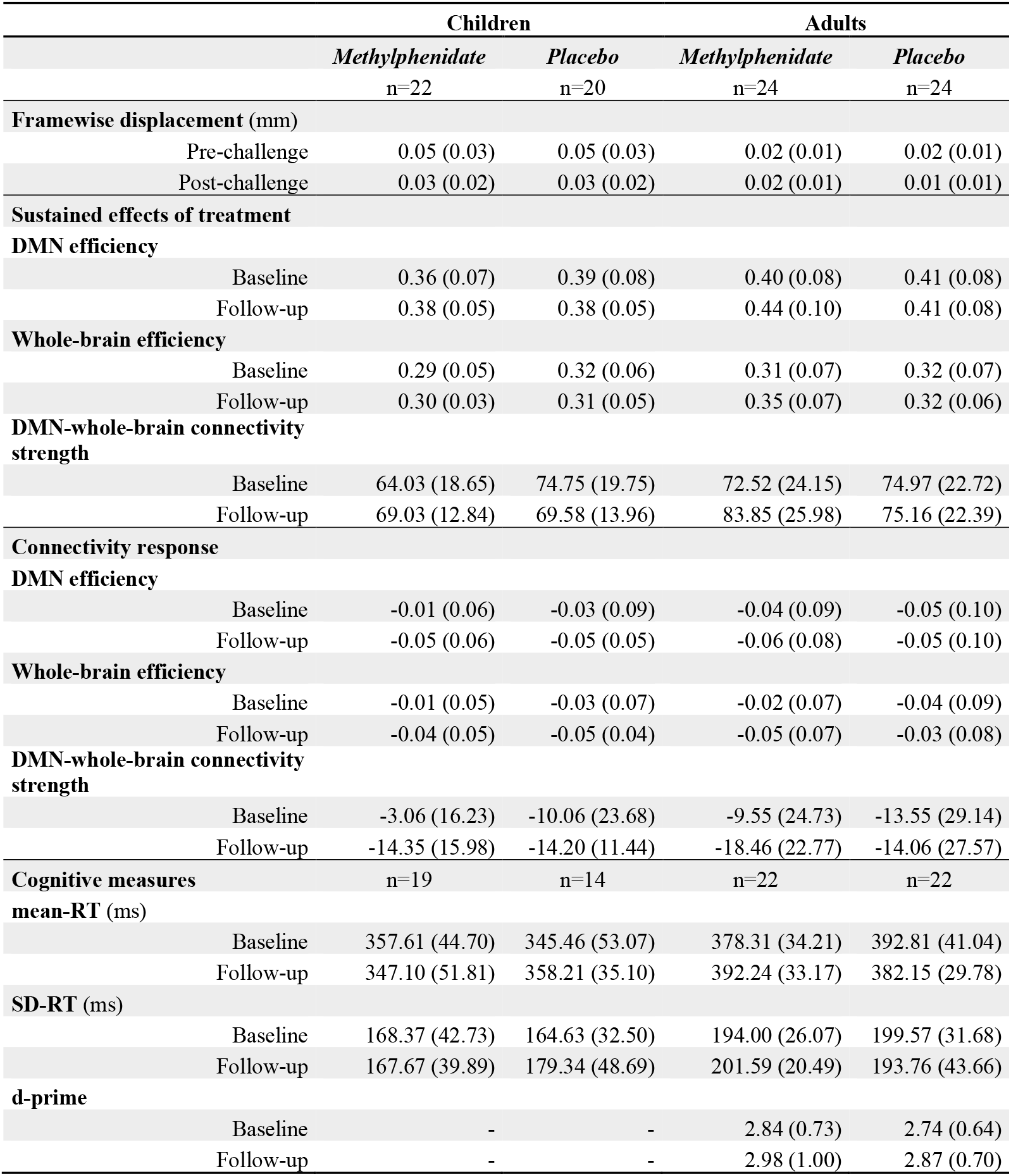
Estimated marginal means of the network analyses in mean (SE). the mean (SD) is summarized for the cognitive measures. DMN = default mode network, RT=reaction time.

### Sustained effects of methylphenidate treatment on functional connectivity

Data are shown in Fig 2 and Supplementary Fig S1. Estimated marginal means are summarized in Table 2.

**Fig 2.**
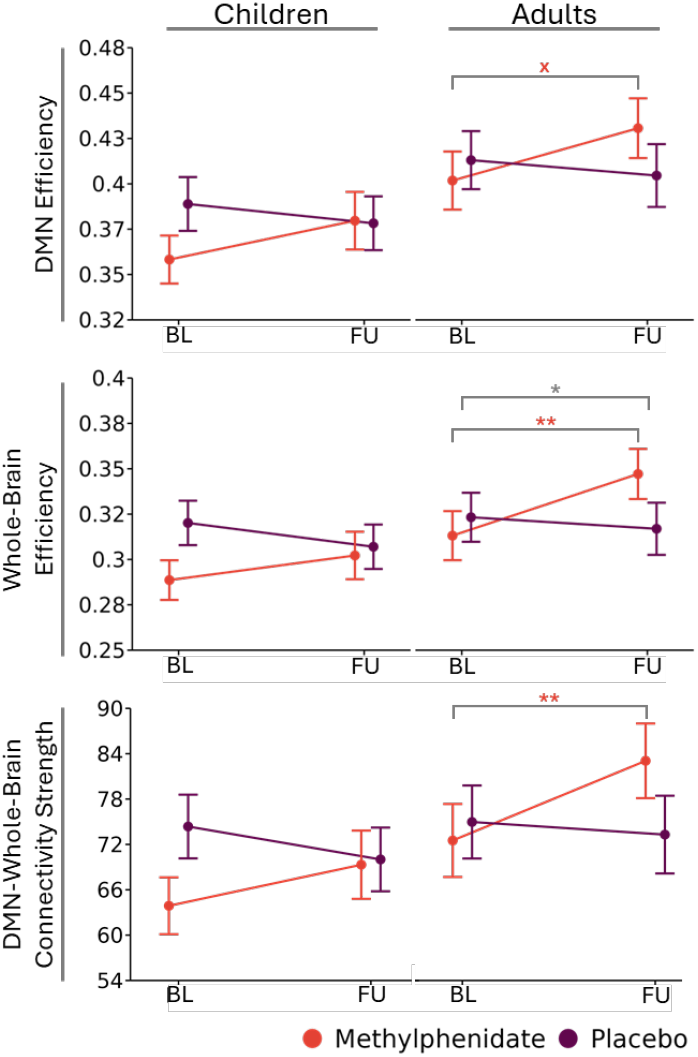
Sustained alterations in functional connectivity following treatment. Line plots represent the estimated marginal means (± SE) of each network measure at baseline and at follow-up for each age and treatment group. DMN = default mode network, BL = baseline, FU = follow-up, *** = p<0.0001, ** = p<0.001, * = p<0.05, x = p<0.1

#### DMN efficiency

While we found no significant main or interaction effects of treatment and visit in adults (all p’s>0.08), a trend increase in DMN efficiency from baseline to follow-up was observed in the methylphenidate-treated group (p=0.06), but not the placebo-treated group (p=0.58). In children, we found no significant main or interaction effects of treatment and visit (all p’s>0.13).

#### Whole-brain efficiency

In adults, we found a significant main effect of visit (χ^2^(1)=3.88,p<0.05) and a treatment-by-visit interaction (χ^2^(1)=6.88, p<0.01). Post-hoc evaluation revealed a trend increase in global efficiency from baseline to follow-up (p=0.08), which was driven by a significant increase in the methylphenidate group (p<0.01), but not the placebo group (p=0.58). In contrast, in children we found no significant main or interaction effects of treatment and visit (all p’s>0.13).

#### DMN-whole-brain connectivity strength

In line with the findings for whole-brain efficiency, in adults we found a significant treatment-by-visit interaction for DMN-whole-brain connectivity strength (χ^2^(1)=4.81, p=0.03). Post-hoc evaluation revealed that DMN-whole-brain connectivity strength increased significantly in the methylphenidate-treated group (p<0.01), but not in the placebo-treated group (p=0.68). No further significant effects were found (all p’s>0.08). In children, we found no significant main or interaction effects of visit and treatment on DMN-whole-brain connectivity strength (all p’s>0.17).

### Development of tolerance: changes in connectivity responses

Data are shown in Fig 3 and Supplementary Fig S2. Estimated marginal means are summarized in Table 2.

**Fig 3.**
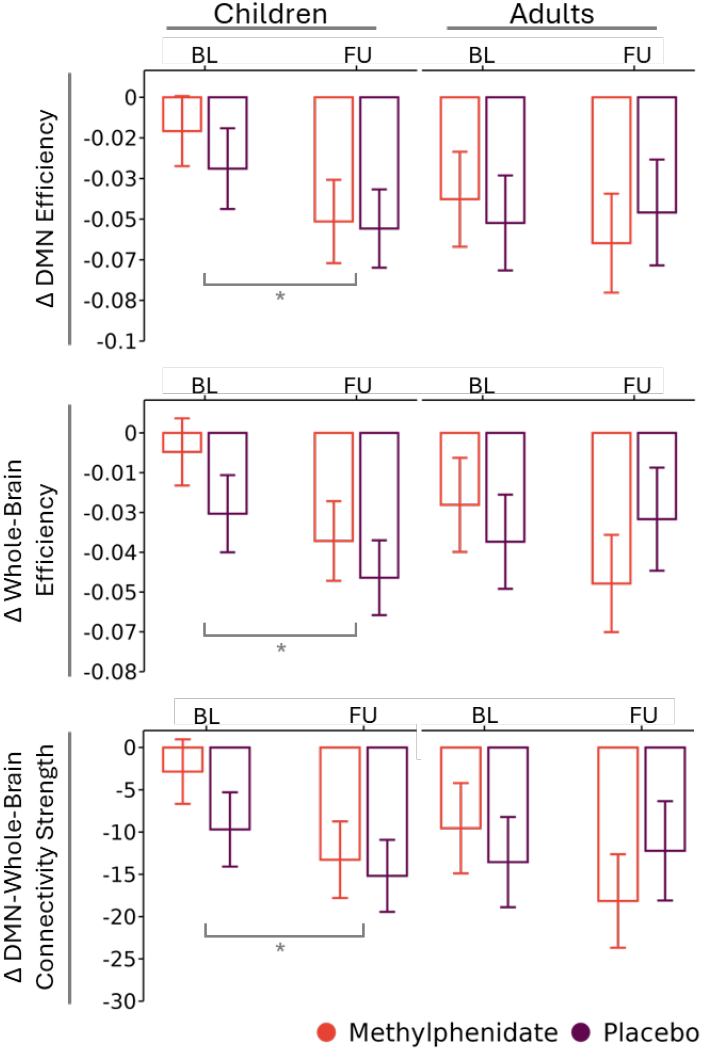
Changes in connectivity responses to a methylphenidate challenge. The bar plots show the estimated marginal means (± SE) response to the methylphenidate-challenge for each age and treatment group for each visit. DMN = default mode network, BL = baseline, FU = follow-up, *** = p<0.0001, ** = p<0.001, * = p<0.05, x = p<0.1

#### DMN efficiency

In adults, no significant main or interaction effects of treatment or visits were found on DMN efficiency response, indicating that the acute challenge elicited similar changes in DMN efficiency at baseline and follow-up (all p’s>0.50).

Pairwise analyses revealed that methylphenidate induced a significant decrease in DMN efficiency (i.e., a significant DMN efficiency response) both at baseline (t_(47)_=-3.32, p=0.001) and follow-up (t_(40)_=-3.95, p<0.001).

In children, we found a main effect of visit on DMN efficiency response (χ^2^(1)=4.98, p=0.026), reflecting a more pronounced decrease in DMN efficiency at follow-up than at baseline across treatment groups (p=0.04). No further main or interactions emerged in children (all p’s>0.58).

Pairwise analyses showed a trend decrease in DMN efficiency following the methylphenidate challenge at baseline (t_(34)_=-1.85, p=0.07) and a significant decrease in DMN efficiency at follow-up(t_(29)_=-5.13, p<0.0001).

#### Whole-brain efficiency

In adults, we found no significant main or interaction effects of treatment or visit on whole-brain efficiency response (all p’s>0.15), suggesting that the acute challenge elicits similar changes in whole-brain efficiency before and after the treatment period.

Pairwise analyses showed that DMN-whole-brain connectivity strength did not decrease significantly after the methylphenidate-challenge at baseline (t_(34)_=-1.82, p=0.07), but it did at follow-up (t_(29)_=-5.79, p<0.0001).

Pairwise analyses illustrated that whole-brain efficiency decreased significantly following the methylphenidate-challenge, both at baseline (t_(47)_=-2.62, p=0.01) and follow-up (t_(40)_=-3.88, p<0.001).

In children, we did find a main effect of visit on whole-brain efficiency response to the methylphenidate-challenge (χ^2^(1)=5.84, p=0.015). More specifically, whole-brain efficiency reduced significantly following the methylphenidate challenge follow-up (p=0.02) in both the methylphenidate and placebo conditions. No further main or interaction effects were found in children (all p’s>0.23).

Pairwise analyses revealed no significant changes in whole-brain efficiency following the methylphenidate challenge at baseline (t_(34)_=-1.67, p=0.10). However, at follow-up, whole-brain efficiency significantly decreased following the challenge (t_(29)_=-5.7185, p<0.0001).

#### DMN-whole-brain connectivity strength

In adults, we found no significant changes in DMN-whole-brain connectivity strength in response to an acute methylphenidate challenge in adults (all p’s>0.14), suggesting that the treatment period had no effect on the acute challenge effects in adults.

We found significant decreases in DMN-whole-brain connectivity strength after the methylphenidate-challenge both at baseline (z_(47)_=-2.98, p<0.01) and follow-up (t_(40)_=-4.23, p=0.0001).

In children, we found a main effect of visit (χ^2^(1)=4.89, p=0.027). Specifically, a larger decrease in DMN-whole-brain connectivity strength following the methylphenidate-challenge was found at follow-up compared to baseline across treatment conditions (p=0.04). We found no further main or interaction effects in children (all p’s>0.33).

### Association of cognitive measures with whole-brain efficiency

Data are shown in Table 2 and Supplementary Fig S3 and S4.

In adults, we found no significant main or interaction effects of treatment group and whole-brain efficiency changes on changes in cognitive measures (p’s> 0.11). The model without d-prime showed a trend-significant interaction effect of treatment group and whole-brain efficiency (F(2,33)=2.89, p=0.07). Post-hoc correlation analyses revealed a positive association between the change in inverse mean-RT and change in whole brain efficiency in the placebo condition (t(15)=2.26, p=0.04, r=0.50), but not the methylphenidate condition (t(19)=-0.726, p=0.48, r=-0.16). No other associations were observed.

In children, we found a significant interaction effect of treatment group and whole-brain efficiency changes on change in cognitive measures (F(2,16)=8.26, p<0.01). Post-hoc correlation analyses revealed a positive association between changes in whole-brain efficiency and changes in inverse mean-RT (t(8)= 2.44, p=0.04, r=0.65), but not inverse SD-RT (t(8)=0.01, p=0.99, r<0.01) in the placebo condition. In the methylphenidate condition, a negative association was found between changes in whole-brain efficiency and changes in inverse mean-RT (t(9)=-3.36, p<0.01, r=-0.75) and inverse SD-RT (t(9)=-1.81, p=0.10, r=-0.52).

## Discussion

This study examined the sustained effects of a 4-month methylphenidate treatment on functional connectivity and on the connectivity response to a methylphenidate-challenge in male children and adults with ADHD to investigate the development of tolerance. We found increased whole-brain efficiency and DMN-whole-brain connectivity strength in adults treated with methylphenidate, but not in those receiving a placebo. Following an acute dose of methylphenidate, global efficiency and connectivity strength decreased. This response remained consistent after a 4-months treatment in adults, and this decrease became larger in children, regardless of medication group. Taken together, the current results suggest that tolerance to the brain’s acute response to methylphenidate does not develop over a 4-month period, in children or adults.

### Sustained effects of a 4-month methylphenidate treatment on network measures

We found sustained increases in whole-brain efficiency and DMN-whole-brain connectivity in methylphenidate-treated adults, but not in placebo-treated adults. A similar, non-significant trend was seen in children, with increases only in the methylphenidate group. Childhood and early adolescence are characterized by large neuronal changes (e.g. pruning) and heightened synaptic plasticity[36]. Preclinical studies investigating the effects of early methylphenidate treatment on neuron function suggests that the juvenile prefrontal cortex is particularly to the effects of methylphenidate[11,37,38], making the absence of significant effects in children unexpected. The absence of significant effects in children may be explained by higher within-subject variability, which could have resulted from higher motion levels during the scans. Overall, our results do not strongly support age-dependent effects of methylphenidate on these network measures. Rather, these findings suggest that methylphenidate treatment induces large-scale changes in brain network topology, persisting up to one week beyond treatment cessation. We cannot rule out that these changes stem from acute discontinuation effects, such as a rebound or compensatory mechanisms, as we lack imaging data from the final treatment day.

Increases in global efficiency are suggested to reflect improvements in information exchange across remote brain regions[33], and high global efficiency has been linked to higher global intelligence and improved performance on working memory tasks[39]. Conversely, lower measures of global efficiency are typically reported in ADHD populations[40]. However, in absence of a neurotypical control sample, we cannot conclude whether the global efficiency increases reflect changes aligning with more neurotypical states.

While we found no significant changes in within-DMN efficiency over time, connectivity strength between the DMN and other brain regions significantly increased following the methylphenidate treatment. This suggests that the importance of the DMN within the whole-brain network increases, while the within-network efficiency of the DMN does not significantly change. Increased DMN connectivity with other resting-state networks has been systematically observed in ADHD populations, and has also been associated with suppression of goal-directed cognition[20,41]. Additionally, prior research found increased DMN-putamen connectivity following a 6-month methylphenidate treatment[26]. Taken together, our findings suggest that prolonged methylphenidate treatment might induce lasting increases in DMN connectivity, however, more research is needed to investigate the link between increased DMN connectivity and changes in cognitive performance. Our exploratory analyses showed that increases in global efficiency were linked to improvements in reaction times of placebo-treated children and adults. In contrast, methylphenidate-treated children showed the opposite pattern: increases in global efficiency were associated with worsening reaction time task performance, suggesting a moderating effect of methylphenidate treatment on this relationship. Interestingly, negative associations between global efficiency and reaction time have been found, with DMN connectivity moderating this relationship[41]. One possible explanation is that highly integrated brains may not be able to further increase network efficiency during task engagement[41]. However, these results should be interpreted with caution, considering the sample size included in the behavioral analyses was limited.

### Development of tolerance

In contrast to the sustained increases, the acute methylphenidate-challenge resulted in significant decreases in all network measures, across both visits, following the challenge in adults, and at the follow-up session in children, irrespective of treatment group. Our results are in line with prior studies that reported similar decreases in functional connectivity. For instance, two studies reported decreases in functional connectivity between the DMN and other brain networks, including the visual network, following a methylphenidate-challenge[21,42]. DMN activity is typically suppressed during task states, and impaired DMN suppression is associated with ADHD pathology and cognitive impairments[20,41]. Decreases in global efficiency following methylphenidate administration may reflect reduced integration between brain regions that do not contribute to performance during task states, e.g., less interference of brain regions during task states.

Contrary to our hypothesis, we found no evidence for the development of tolerance in our chosen parameters. In adults, connectivity responses remained stable from baseline to follow-up across all network measures. This finding contrasts with prior reports of both tolerance and sensitization following prolonged methylphenidate treatment, likely due to methodological differences such as age-at-treatment start, treatment duration, dosage, and outcome measures[17,18,43]. Notably, tolerance is more commonly observed in studies with longer treatment durations and higher doses[2,16], whereas studies with shorter treatment durations tend to report sensitization[18]. Our treatment duration of four months was relatively short in comparison to studies that reported behavioral tolerance, which typically spanned treatment periods of at least a year[2,3,7]. Therefore, it may be possible that tolerance develops over longer treatment periods. Moreover, a meta-analysis of the treatment efficacy of methylphenidate reported that efficacy loss increased with study duration[44]. Lastly, the one-week washout period in our study may have allowed any potential tolerance effects to recover, as acute behavioral tolerance has been shown not to carry over to the next day[15].

To our surprise, we observed a significant larger decrease in connectivity response in children, irrespective of treatment condition. Since these changes were not specific to methylphenidate treatment, these changes are not due to sensitization. Instead, they may reflect underlying developmental processes or non-pharmacological treatment effects.

Parents of both groups received psychoeducation about ADHD and access to a treatment provider, which may have, in turn, influenced the neural response to methylphenidate. While it is possible that decreases in motion between pre- and post-challenge scans influenced our findings on the acute effects of methylphenidate, we only found small, trend-significant decreases in motion from baseline to follow-up in children.

### Strengths, limitations, future directions

The main strengths of this study are the inclusion of medication-naïve participants and randomized allocation to methylphenidate or placebo, allowing developmental effects to be distinguished from medication effects. However, the study has some limitations. Only male participants were included, despite ADHD and stimulant treatment effects remaining critically understudied in females[25]. Sex-related differences have been reported in both ADHD symptom presentation[31] and treatment response[46], limiting the generalizability of our findings to the full ADHD population. Additionally, the 4-month treatment period may have been too short to detect tolerance, which typically develops over several years — though ethical constraints make longer RCTs challenging. Future preclinical studies can complement this work by offering greater experimental control and invasive techniques to investigate the multi-scale mechanisms underlying methylphenidate’s effects.

## Data availability

The data supporting the conclusions of this study are available from the corresponding author upon reasonable request.

## Acknowledgements

We would like to thank the participants of the effects of Psychotropic drugs On the Developing brain (ePOD-MPH) study for their participation in the study.

## Funding

This study was funded by the ZonMW open grant (project number: 09120012110091). The ePOD-MPH trial was funded by a research grant awarded to LR by the Academic Medical Center, University of Amsterdam, and 11.32050.26ERA-NET PRIOMEDCHILD FP 6(EU). JC received funding from the NIH grants R01MH119091 and R01MH136041.

## Ethics Declaration Ethics approval

The study is in compliance with the code of medical ethics, and was approved on March 24, 2011 by the Central Committee on Research Involving Human Subjects (NL34509.000.10), and was registered at under trial number NTR3103 with The Netherlands National Trial Register.

## Consent to participate

All participants and parents or legal representatives of the children provided written informed consent.

## Accordance with consort guidelines

The completed CONSORT checklist can be found in supplementary information.

## Competing interests

The authors declare no conflicts of interest.

## Notes

### Competing Interest Statement

The authors have declared no competing interest.

### Clinical Trial

NTR3103

### Author Declarations

The study was approved on March 24, 2011 by the Central Committee on Research Involving Human Subjects (NL34509.000.10)

### Summary of Updates

The authors further expanded on the research gaps that the manuscript aims to bridge, adjusted tables for legibility, clarified statistical testing approaches including multiple comparison correction approach.

