## Supplemental Material for "Effects of 4-Month Methylphenidate Treatment on Functional Connectivity in Male Individuals with Attention-Deficit Hyperactivity Disorder"

### Supplementary methods

#### Participants

All participants were diagnosed with ADHD (DSM-IV, all types) by an experienced psychiatrist, using a structured interview (Diagnostic Interview Schedule for Children (NIMH-DISC-IV): authorized Dutch translation[1] and the Diagnostic Interview for ADHD (DIVA 2.0) for adults[2]. Participants were included if at least 6 inattention or hyperactivity/impulsivity symptoms according to the DSM-IV were present. Participants were not eligible for inclusion if they had received prior or current clinical treatments, or had prior or current dependencies on drugs influencing the dopaminergic system, and in adults, if the treatments took place before age 23. Additional exclusion criteria included having an estimated IQ < 80, assessed using the Block Design and Vocabulary subtests of the Wechsler Intelligence Scale for Children - Revised (WISC-III-R)[3] in children, and the Dutch Adult Reading[4] in adults. Lastly, participants were excluded if they had a history of significant medical or neurological trauma or illness.

#### Methylphenidate treatment

Participants received a placebo or oral dosages of short-acting MPH, starting with 1-2 doses of 0.3 mg/kg daily, and dosages were increased weekly with 5-10 mg/day until the clinical target dosage was reached according to the clinical guidelines of the Netherlands, with a maximum of 50 mg/day. If serious side-effects occurred after increasing or decreasing a dosage, the participant returned to the previous dosage, and dosage modifications were more gradual thereafter. All participants, including the placebo group received all other active components of treatment during the trial, including thorough evaluation, psycho-education about ADHD, access to a treatment provider, supportive care, and the expectation of improvement.

#### Randomization, treatment allocation and blinding

Randomization was performed using a specialized computer program developed by the Clinical Research Unit of the AMC. Participants were stratified by age and randomized to either placebo or methylphenidate treatment using a permuted block randomization scheme. Subjects were randomized after the first MRI scan at baseline. Treatment allocation was concealed for all parties. Blinding was done by the Clinical Research Unit of the AMC, methylphenidate and placebo were provided in identical form in 10mg tablets, in similar container types, dispensing aids and formulation number. After the study end, blinding was checked with the patient, the treating psychiatrist and the study investigators.

### Sample size calculation

To detect a standardized effect size of 1.25 with a two-sided significance level of 5%, a sample size of 15 patients in each group (4 groups, 2 treatment groups x 2 age groups) was calculated as sufficient. Considering a 25% drop-out rate, the final included sample size was 25 participants per group.

### Cognitive and behavioral assessments

#### *ADHD symptom severity*

ADHD symptom severity was assessed at baseline and follow-up, using the Inattentive Symptoms and Hyperactivity/Impulsive Symptoms subscales of the Disruptive Behavior Disorder Rating-Scale (DBD-RS)[5] in children, and the ADHD-Self Report (ADHD-SR)[2] total score in adults.

#### *Go/no-go task*

In addition to the resting-state fMRI scans, an adaptation of the go/no-go task[6] was administered during MRI scanning to assess psychomotor inhibition. During go trials, the participant was instructed to press a button when they were presented a target stimulus (a Pokémon cartoon character). During no-go trials, the non-target stimulus was presented, and the participants were instructed not to press the button. Stimulus presentation lasted 500 ms, and the inter-stimulus-interval (ISI) was 3000 ms. The task consisted of 3 runs, each containing 57 trials (43 Go-trials and 14 No-Go trials), in pseudo-randomized order of no-go trials. The outcome variable was *d-prime*[7], and is calculated as the difference between the proportion of correct go-trials and the proportion of incorrect no-go trials. Higher d-prime values reflect better psychomotor inhibition than lower d-prime values.

#### *Reaction Time (RT) task*

To assess psychomotor speed, a reaction speed test was designed based on the Amsterdam Neuropsychological Tasks (ANT) battery[8]. Participants were instructed to press a button as fast as possible when they were presented with a target stimulus on the center of the screen (a [9]friendly-looking monster). The test consisted of 12 practice trials, after which 30 stimuli were presented for each hand, with an ISI between 500 – 1500 ms. Hand-order was counterbalanced, and only performance with the participants' dominant hand was included in analysis. The outcome variables were mean and standard deviation (sd) of the reaction time, as ADHD is associated with slower (high means) and more variable (high sd) RT's[9].

### MRI acquisition

The MRI scans were acquired on two 3T Philips scanners ( Achieva / Intera, Philips Healthcare, Best, The Netherlands), using an 8-channel receive-only head coil. The total resting-state scan duration was 4 min and 58 sec. Participants were instructed to keep their eyes open and let their mind wander. To increase compliance, light blue blocks were shown on a white background, disappearing one by one every minute to indicate the duration of the scan. The screen was placed directly behind the scanner and participants were able to view the screen through a mirror system attached to the head coil. Eight adult participants prematurely received their follow-up scan after 9 weeks due to scheduled scanner maintenance.

### MRI preprocessing

MRI Preprocessing was performed using FMRIPREP v1.2.3 for all but one scan, which was processed using FMRIPREP V20.1.1 due to registration issues. Each T1w image was bias-corrected, skull stripped, and normalized to the MNI space using non-linear registration. Functional data preprocessing included motion correction using FLIRT. This was followed by co-registration to the corresponding T1w using boundary-based registration with 9 degrees of freedom.

Motion correcting transformations, BOLD-to-T1w transformation, and T1w-to-template (MNI) warp were concatenated and applied in a single step using antsApplyTransforms (ANTs v2.1.0) with Lanczos interpolation. Independent component analysis (ICA) based on Automatic Removal Of Motion Artifacts (AROMA) was used to generate data that was non-aggressively denoised[10]. Two brain volumes were removed from the start of each scan to ensure that the steady-state equilibrium was attained.

High frequency motion was quantified by determining the percent of relative power above 0.1 Hz within each motion direction, focusing on motion in the y-translation (phase-encoding), using scripts and protocols from Gratton et al (2020)[11], using a low-pass Butterworth filter of 1st order, with normalized cutoff frequency 0.1/(0.5/TR) with TR=2.3

Temporal signal-to-noise (tSNR) maps were calculated per participant, and a mask was made to exclude the voxels in the 25th lowest tSNR percentile. Regions were bilaterally excluded from analysis if more than 70% of voxels were excluded due to low tSNR, in at least 90% of participants. In supplementary table 1, an overview of the excluded regions is given. Region 233, part of the pre-motor thalamus was entirely excluded by the gray matter mask, and was bilaterally excluded in addition to the low-tSNR regions.

**R statistical tests**

Statistical models were constructed as follows:
global_efficiency ~ visit * treatment + (1|participant)
car::Anova(model)

**Sensitivity analyses**

To assess the relation between motion and the independent variables, LMM's were constructed separately for each age group (children, adults), with visit (baseline, follow-up), methylphenidate challenge (pre-challenge, post-challenge) and treatment group (methylphenidate, placebo) as independent variables, and mean FD during each scan as the outcome measure.

*Table S1 Excluded low-tSNR BNA regions*

| Lobe | Gyrus | Left and right Hemisphere | Label ID.L | Label ID.R | Anatomical and modified Cyto-architectonic descriptions | lh.MNI(X,Y,Z) | rh.MNI(X,Y,Z) |
|---|---|---|---|---|---|---|---|
| **Frontal Lobe** | OrG, Orbital Gyrus | OrG_L(R)_6_4 | 47 | 48 | *A11m, medial area 11* | -6, 52, -19 | 6, 57, -16 |
| | | OrG_L(R)_6_5 | 49 | 50 | *A13, area 13* | -10, 18, -19 | 9, 20, -19 |
| **Temporal Lobe** | STG, Superior Temporal Gyrus | STG_L(R)_6_1 | 69 | 70 | *A38m, medial area 38* | -32, 14, -34 | 31, 15, -34 |
| | MTG, Middle Temporal Gyrus | MTG_L(R)_4_1 | 81 | 82 | *A21c, caudal area 21* | -65, -30, -12 | 65, -29, -13 |
| | ITG, Inferior Temporal Gyrus | ITG_L(R)_7_1 | 89 | 90 | *A20iv, intermediate ventral area 20* | -45, -26, -27 | 46, -14, -33 |
| | | ITG_L(R)_7_3 | 93 | 94 | *A20r, rostral area 20* | -43, -2, -41 | 40, 0, -43 |
| | | ITG_L(R)_7_4 | 95 | 96 | *A20il, intermediate lateral area 20* | -56, -16, -28 | 55, -11, -32 |
| | | ITG_L(R)_7_7 | 101 | 102 | *A20cv, caudoventral of area 20* | -55, -31, -27 | 54, -31, -26 |
| | PhG, Parahippocampal Gyrus | PhG_L(R)_6_1 | 109 | 110 | *A35/36r, rostral area 35/36* | -27, -7, -34 | 28, -8, -33 |
| | | PhG_L(R)_6_2 | 111 | 112 | *A35/36c, caudal area 35/36* | -25, -25, -26 | 26, -23, -27 |
| | | PhG_L(R)_6_4 | 115 | 116 | *A28/34, area 28/34 (EC, entorhinal cortex)* | -19, -12, -30 | 19, -10, -30 |
| | | PhG_L(R)_6_5 | 117 | 118 | *TI, area TI(temporal agranular insular cortex)* | -23, 2, -32 | 22, 1, -36 |
| **Occipital Lobe** | MVOcC, MedioVentral Occipital Cortex | MVOcC _L(R)_5_1 | 189 | 190 | *cLinG, caudal lingual gyrus* | -11, -82, -11 | 10, -85, -9 |
| | | MVOcC _L(R)_5_2 | 191 | 192 | *rCunG, rostral cuneus gyrus* | -5, -81, 10 | 7, -76, 11 |
| | | MVOcC _L(R)_5_3 | 193 | 194 | *cCunG, caudal cuneus gyrus* | -6, -94, 1 | 8, -90, 12 |
| | LOcC, lateral Occipital Cortex | LOcC _L(R)_4_3 | 203 | 204 | *OPC, occipital polar cortex* | -18, -99, 2 | 22, -97, 4 |
| **Subcortical Nuclei** | Tha, Thalamus | Tha_L(R)_8_2 | 233 | 234 | *mPMtha, pre-motor thalamus* | -18, -13, 3 | 12, -14, 1 |

*Table S2 Default mode network BNA atlas parcellation numbers*

| Lobe | Gyrus | Left and Right Hemisphere | Label ID.L | Label ID.R | Anatomical and modified Cyto-architectonic descriptions | lh.MNI(X,Y,Z) | rh.MNI(X,Y,Z) |
|---|---|---|---|---|---|---|---|
| Frontal Lobe | SFG, Superior Frontal Gyrus | SFG_L(R)_7_2 | 3 | 4 | *A8dl, dorsolateral area 8* | -18, 24, 53 | 22, 26, 51 |
| | | SFG_L(R)_7_3 | 5 | 6 | *A9l, lateral area 9* | -11, 49, 40 | 13, 48, 40 |
| | | SFG_L(R)_7_6 | 11 | 12 | *A9m,medial area 9* | -5, 36, 38 | 6, 38, 35 |
| | | SFG_L(R)_7_7 | 13 | 14 | *A10m, medial area 10* | -8, 56, 15 | 8, 58, 13 |
| | MFG, Middle Frontal Gyrus | MFG_L(R)_7_5 | 23 | 24 | *A8vl, ventrolateral area 8* | -33, 23, 45 | 42, 27, 39 |
| | | MFG_L(R)_7_7 | 27 | 28 | *A10l, lateral area10* | -26, 60, -6 | 25, 61, -4 |
| | IFG, Inferior Frontal Gyrus | IFG_L(R)_6_3 | 33 | 34 | *A45c, caudal area 45* | -53, 23, 11 | 54, 24, 12 |
| | | IFG_L(R)_6_4 | 35 | 36 | *A45r, rostral area 45* | -49, 36, -3 | 51, 36, -1 |
| | | IFG_L(R)_6_6 | 39 | 40 | *A44v, ventral area 44* | -52, 13, 6 | 54, 14, 11 |
| | OrG, Orbital Gyrus | OrG_L(R)_6_1 | 41 | 42 | *A14m, medial area 14* | -7, 54, -7 | 6, 47, -7 |
| | | OrG_L(R)_6_2 | 43 | 44 | *A12/47o, orbital area 12/47* | -36, 33, -16 | 40, 39, -14 |
| | | OrG_L(R)_6_6 | 51 | 52 | *A12/47l, lateral area 12/47* | -41, 32, -9 | 42, 31, -9 |
| Temporal Lobe | STG, Superior Temporal Gyrus | STG_L(R)_6_6 | 79 | 80 | *A22r, rostral area 22* | -55, -3, -10 | 56, -12, -5 |
| | MTG, Middle Temporal Gyrus | MTG_L(R)_4_1 | 81 | 82 | *A21c, caudal area 21* | -65, -30, -12 | 65, -29, -13 |
| | | MTG_L(R)_4_2 | 83 | 84 | *A21r, rostral area 21* | -53, 2, -30 | 51, 6, -32 |
| | | MTG_L(R)_4_4 | 87 | 88 | *aSTS, anterior superior temporal sulcus* | -58, -20, -9 | 58, -16, -10 |
| | ITG, Inferior Temporal Gyrus | ITG_L(R)_7_4 | 95 | 96 | *A20il, intermediate lateral area 20* | -56, -16, -28 | 55, -11, -32 |
| | PhG, Parahippocampal Gyrus | PhG_L(R)_6_3 | 113 | 114 | *TL, area TL (lateral PPHC, posterior parahippocampal gyrus)* | -28, -32, -18 | 30, -30, -18 |
| | pSTS, posterior Superior Temporal Sulcus | pSTS_L(R)_2_1 | 121 | 122 | *rpSTS, rostroposterior superior temporal sulcus* | -54, -40, 4 | 53, -37, 3 |
| | | pSTS_L(R)_2_2 | 123 | 124 | *cpSTS, caudoposterior superior temporal sulcus* | -52, -50, 11 | 57, -40, 12 |
| Parietal Lobe | Pcun, Precuneus | PCun_L(R)_4_3 | 151 | 152 | *dmPOS, dorsomedial parietooccipital sulcus(PEr)* | -12, -67, 25 | 16, -64, 25 |
| | | PCun_L(R)_4_4 | 153 | 154 | *A31, area 31 (Lc1)* | -6, -55, 34 | 6, -54, 35 |
| Insular Lobe | INS, Insular Gyrus | INS_L(R)_6_2 | 165 | 166 | *vIa, ventral agranular insula* | -32, 14, -13 | 33, 14, -13 |
| Limbic Lobe | CG, Cingulate Gyrus | CG_L(R)_7_1 | 175 | 176 | *A23d, dorsal area 23* | -4, -39, 31 | 4, -37, 32 |
| | | CG_L(R)_7_3 | 179 | 180 | *A32p, pregenual area 32* | -6, 34, 21 | 5, 28, 27 |
| | | CG_L(R)_7_4 | 181 | 182 | *A23v, ventral area 23* | -8, -47, 10 | 9, -44, 11 |
| | | CG_L(R)_7_7 | 187 | 188 | *A32sg, subgenual area 32* | -4, 39, -2 | 5, 41, 6 |

**Supplementary results**

**Motion effects**

***The effect of methylphenidate on motion***

In both children ($\chi^2(1)=14.39$, $p<0.001$) and adults $\chi^2(1)=36.99, p<0.0001$), there was a significant main effect of the methylphenidate challenge on motion, reflecting a significant reduction in motion after the acute methylphenidate challenge. For both age groups, no further main or interaction effects emerged from treatment, visit and methylphenidate challenge (all $p>0.13$). Notably, there was a lot of variability in motion during the scans of the children, but less so in adults (table1, participant characteristics).

***Associations between motion and connectivity measures***

Using repeated measures correlations (R, rmcorr), we assessed the association between motion and our primary outcome measures; whole-brain efficiency, DMN efficiency, DMN-whole-brain connectivity strength. Whole-brain efficiency was positively associated with motion in adults ($r=0.38$ $p<0.0001$), but not in children ($r=0.13$, $p=0.16$). Similarly, DMN-whole-brain connectivity strength was positively associated with motion in adults ($r=0.42$, $p<0.0001$), but not in children ($r=0.25$, $p=0.11$), and DMN efficiency was positively correlated with motion in adults ($r=0.39$, $p<0.0001$), but not in children ($r=0.02$, $p=0.85$).

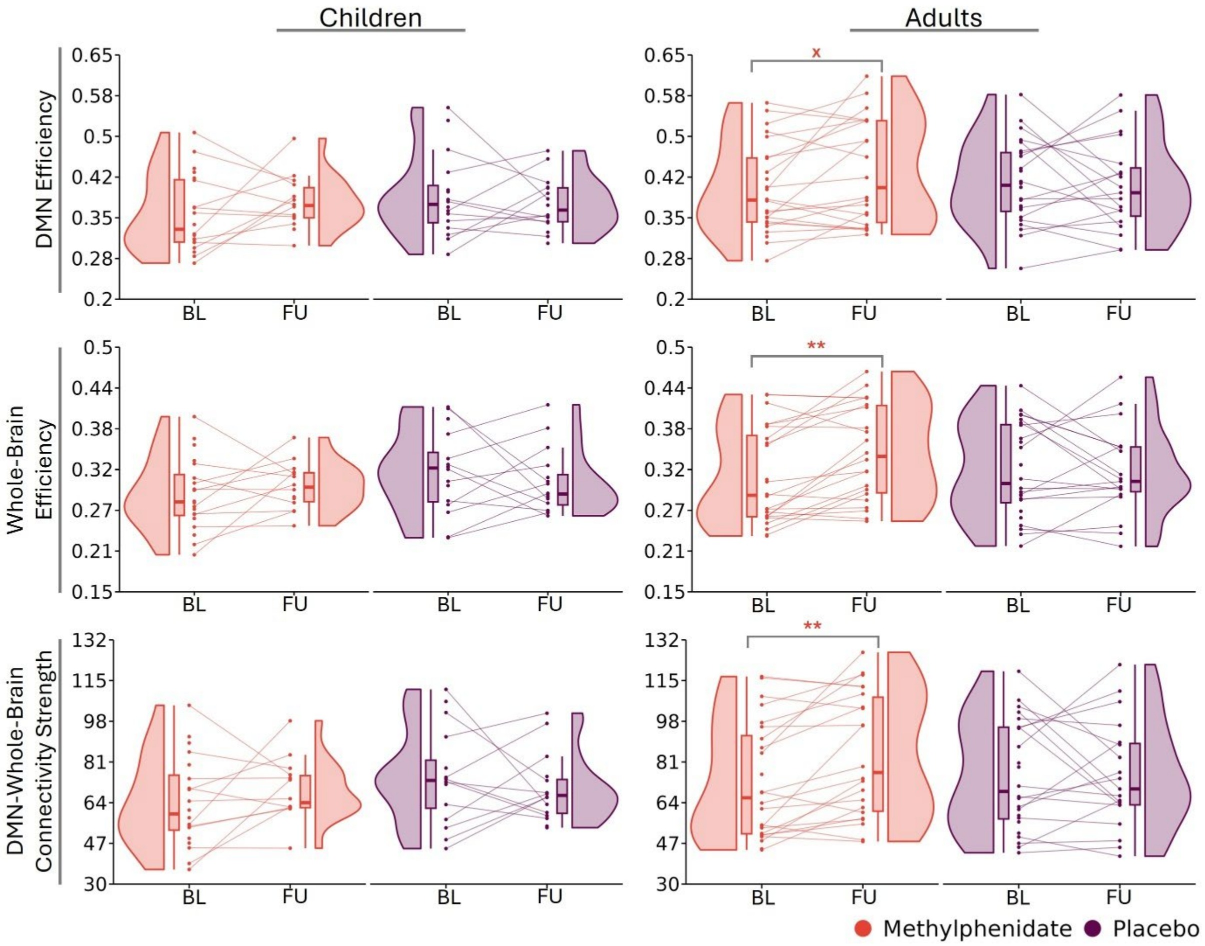


***Supplementary fig S1 Sustained effects of methylphenidate on functional connectivity.*** *Raincloud plots of each connectivity measure at baseline and at follow-up for each age- and treatment group. DMN = default mode network, BL = baseline, FU = follow-up*

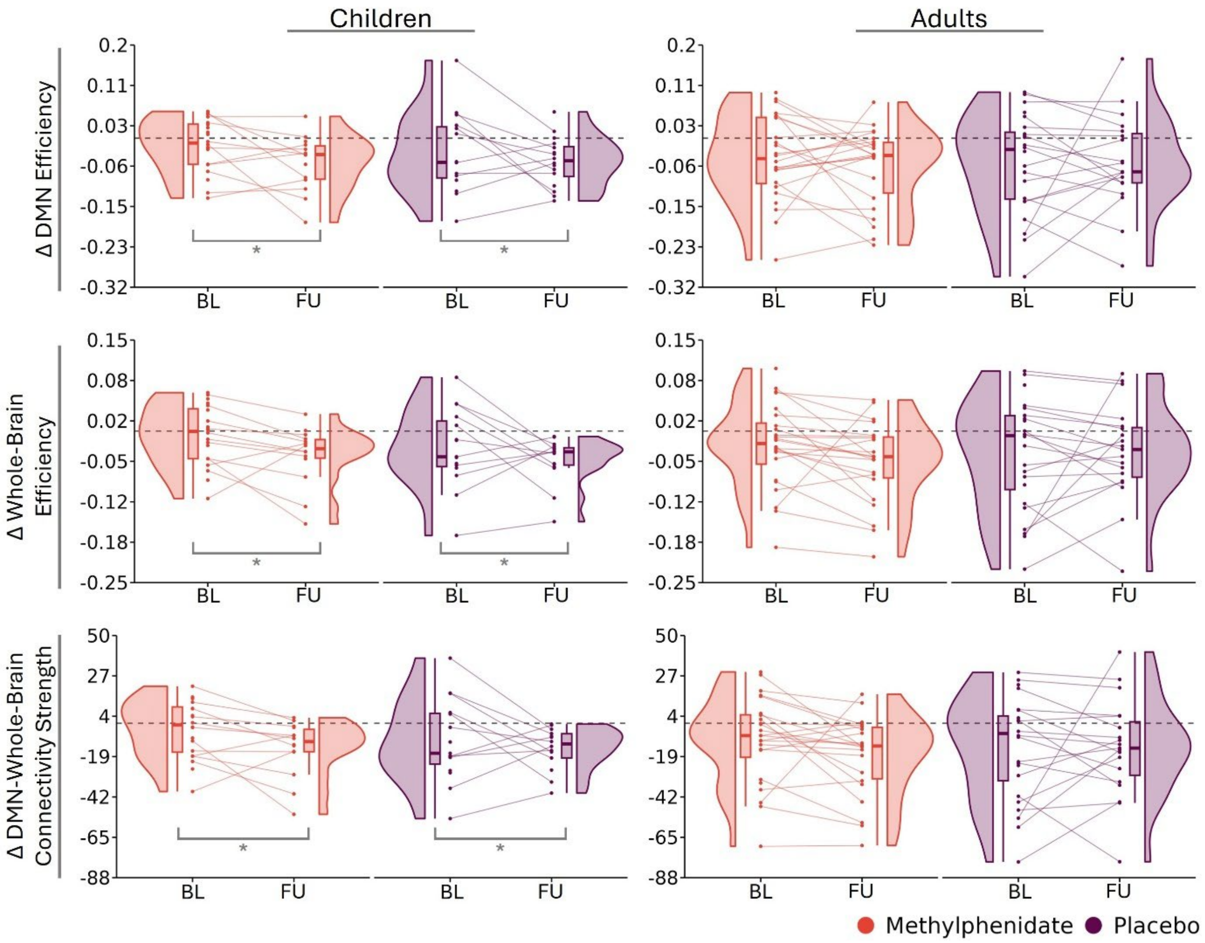


***Supplementary fig 2 Changes in response to a methylphenidate challenge.*** *Raincloud plots of the changes in connectivity response of each connectivity measure at baseline and at follow-up for each age- and treatment group. DMN = default mode network, BL = baseline, FU = follow-up*

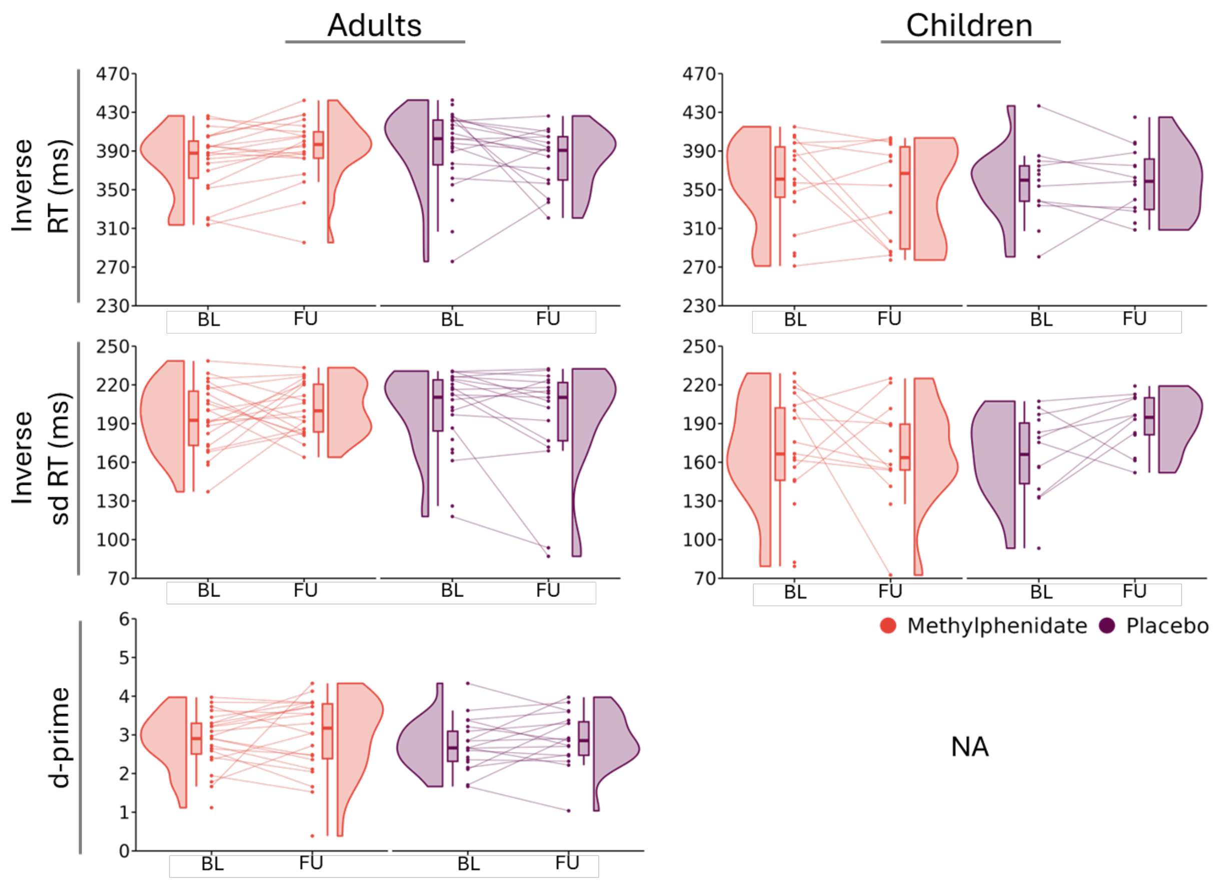


***Supplementary fig S3 Sustained effects of methylphenidate on cognitive outcomes.*** *Raincloud plots of each cognitive measure at baseline and follow-up, separately for ssseach age- and treatment group . RT = mean reaction time, sd RT = reaction time variability*

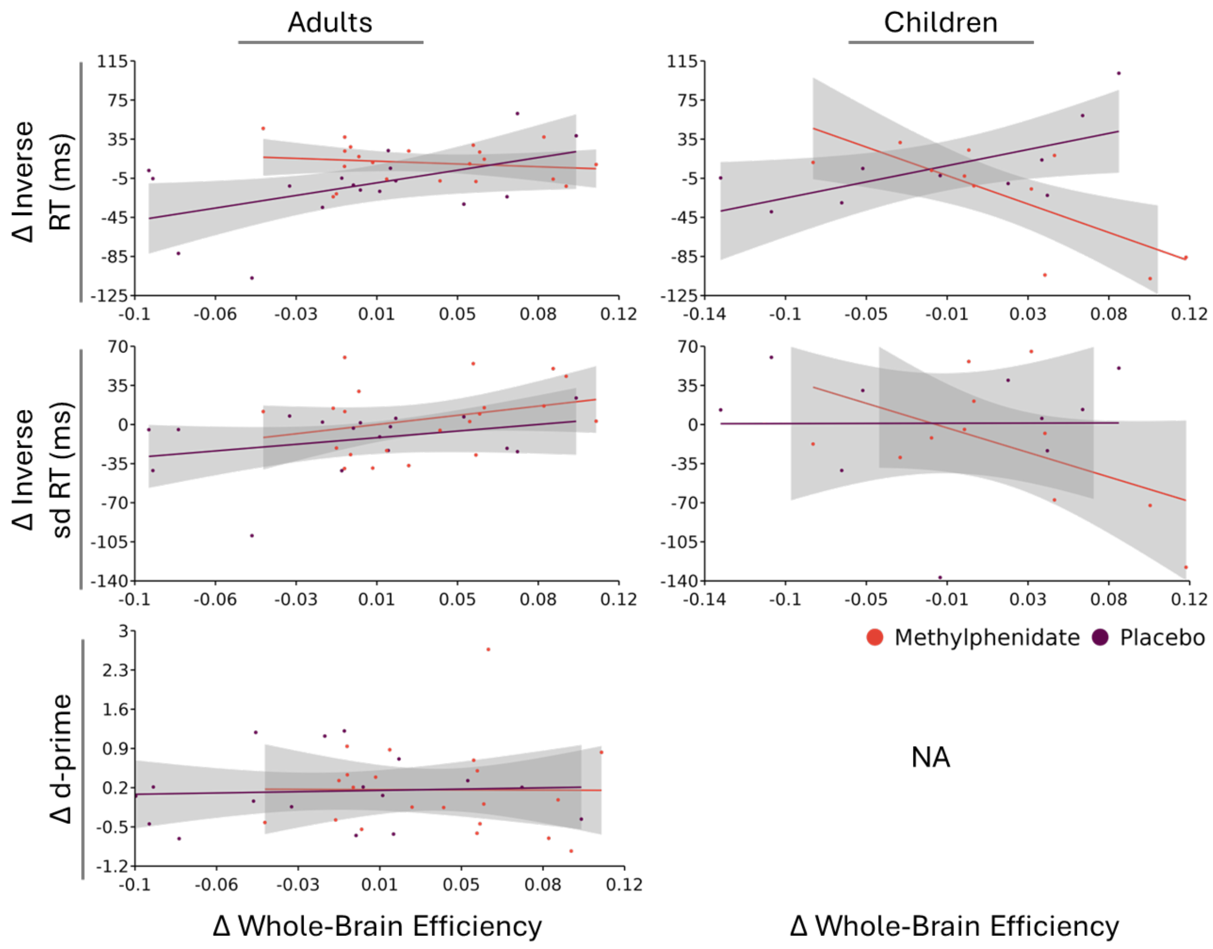


***Supplementary fig S4 Correlations plots of changes in whole-brain efficiency and cognitive measures.*** *Correlation plots of the changes from baseline to follow-up in whole-brain efficiency and changes in the cognitive measures, separately for each age- and treatment group. RT = mean reaction time, sd RT = reaction time variability*

**Table S3 CONSORT 2025 checklist of information to include when reporting a randomised trial**

| Section / Topic | No | CONSORT 2025 checklist item description | Reported on page no. |
|---|---|---|---|
| **Title and abstract** | | | **2** |
| Title and structured abstract | 1a | Identification as a randomised trial | 2,4 |
| | 1b | Structured summary of the trial design, methods, results, and conclusions | 2 |
| **Open science** | | | |
| Trial registration | 2 | Name of trial registry, identifying number (with URL) and date of registration | 4 |
| Protocol and statistical analysis plan | 3 | Where the trial protocol and statistical analysis plan can be accessed | 4 |
| Data sharing | 4 | Where and how the individual de-identified participant data (including data dictionary), statistical code and any other materials can be accessed | 1 |
| Funding and conflicts of interest | 5a | Sources of funding and other support (e.g., supply of drugs), and role of funders in the design, conduct, analysis and reporting of the trial | 1,4 |
| | 5b | Financial and other conflicts of interest of the manuscript authors | 1 |
| **Introduction** | | | |
| Background and rationale | 6 | Scientific background and rationale | 3-4 |
| Objectives | 7 | Specific objectives related to benefits and harms | 3-4 |
| **Methods** | | | |
| Patient and public involvement | 8 | Details of patient or public involvement in the design, conduct and reporting of the trial | N/A |
| Trial design | 9 | Description of trial design including type of trial (e.g., parallel group, crossover), allocation ratio, and framework (e.g., superiority, equivalence, non-inferiority, exploratory) | 4-5 |
| Changes to trial protocol | 10 | Important changes to the trial after it commenced including any outcomes or analyses that were not prespecified, with reason | N/A |
| Trial setting | 11 | Settings (e.g., community, hospital) and locations (e.g., countries, sites) where the trial was conducted | 4, supplements 1 |
| Eligibility criteria | 12a | Eligibility criteria for participants | 4, supplements 1 |
| | 12b | If applicable, eligibility criteria for sites and for individuals delivering the interventions (e.g., surgeons, physiotherapists) | N/A |
| Intervention and comparator | 13 | Intervention and comparator with sufficient details to allow replication. If relevant, where additional materials describing the intervention and comparator (e.g., intervention manual) can be accessed | 4-6 |

| | | | |
|---|---|---|---|
| Outcomes | 14 | Pre-specified primary and secondary outcomes, including the specific measurement variable (e.g., systolic blood pressure), analysis metric (e.g., change from baseline, final value, time to event), method of aggregation (e.g., median, proportion), and time point for each outcome | 5-6 |
| Harms | 15 | How harms were defined and assessed (e.g., systematically, non-systematically) | Supplements 1 |
| Sample size | 16a | How sample size was determined, including all assumptions supporting the sample size calculation | Supplements 1 |
| | 16b | Explanation of any interim analyses and stopping guidelines | N/A |
| Randomisation: | | | |
| Sequence generation | 17a | Who generated the random allocation sequence and the method used | Supplements 1 |
| | 17b | Type of randomisation and details of any restriction (e.g., stratification, blocking and block size) | Supplements 1 |
| Allocation concealment mechanism | 18 | Mechanism used to implement the random allocation sequence (e.g., central computer/telephone; sequentially numbered, opaque, sealed containers), describing any steps to conceal the sequence until interventions were assigned | Supplements 1 |
| Implementation | 19 | Whether the personnel who enrolled and those who assigned participants to the interventions had access to the random allocation sequence | Supplements 1 |
| Blinding | 20a | Who was blinded after assignment to interventions (e.g., participants, care providers, outcome assessors, data analysts) | 5, Supplements 1,2 |
| | 20b | If blinded, how blinding was achieved and description of the similarity of interventions | Supplements 1,2 |
| Statistical methods | 21a | Statistical methods used to compare groups for primary and secondary outcomes, including harms | 6 |
| | 21b | Definition of who is included in each analysis (e.g., all randomised participants), and in which group | 6-7 |
| | 21c | How missing data were handled in the analysis | 6 |
| | 21d | Methods for any additional analyses (e.g., subgroup and sensitivity analyses), distinguishing prespecified from post-hoc | 6 |
| **Results** | | | |
| Participant flow, including flow diagram | 22a | For each group, the numbers of participants who were randomly assigned, received intended intervention, and were analysed for the primary outcome | 6-7 |
| | 22b | For each group, losses and exclusions after randomisation, together with reasons | 6-7 |
| Recruitment | 23a | Dates defining the periods of recruitment and follow-up for outcomes of benefits and harms | N/A |
| | 23b | If relevant, why the trial ended or was stopped | N/A |
| Intervention and comparator delivery | 24a | Intervention and comparator as they were actually administered (e.g., where appropriate, who delivered the intervention/comparator, how participants adhered, whether they were delivered as intended [fidelity]) | 7 |
| | 24b | Concomitant care received during the trial for each group | Supplements 1 |
| Baseline data | 25 | A table showing baseline demographic and clinical characteristics for each group | 7 |

| | | | |
|---|---|---|---|
| Numbers analysed, outcomes and estimation | 26 | For each primary and secondary outcome, by group:<br>● the number of participants included in the analysis<br>● the number of participants with available data at the outcome time point<br>● result for each group, and the estimated effect size and its precision (such as 95% confidence interval)<br>● for binary outcomes, presentation of both absolute and relative effect size | 10 |
| Harms | 27 | All harms or unintended events in each group | N/A |
| Ancillary analyses | 28 | Any other analyses performed, including subgroup and sensitivity analyses, distinguishing pre-specified from post-hoc | 9-10, supplements |
| **Discussion** | | | |
| Interpretation | 29 | Interpretation consistent with results, balancing benefits and harms, and considering other relevant evidence | 11-12 |
| Limitations | 30 | Trial limitations, addressing sources of potential bias, imprecision, generalisability, and, if relevant, multiplicity of analyses | 12 |
